# First time genotyping of *Cryptosporidium* spp. isolates from diarrheic stools of Algerian HIV-Infected subjects supports predominant zoonotic transmission routes

**DOI:** 10.1101/2020.04.14.20055038

**Authors:** Malika Semmani, Damien Costa, Nassima Achour, Meriem Cherchar, Abdelmounaim Mouhajir, Venceslas Villier, Jean Jacques Ballet, Loic Favennec, Haiet Adjmi Hamoudi, Romy Razakandrainibe

## Abstract

**Background:** *Cryptosporidium* is a significant cause of chronic diarrhoea and death in HIV-infected patients. Although HIV-infected patients under HAART have currently reduced risk of suffering from opportunistic infections, opportunistic gastrointestinal infections such as cryptosporidiosis still occur. Currently, there are no data on genetic characteristics of *Cryptosporidium* isolates from cryptosporidiosis patients in Algeria. This study was aimed at identifying *Cryptosporidium* species and subtype families prevalent in Algerian HIV-infected patients and contributing to the molecular epidemiology mapping of *Cryptosporidium* in the MENA region.

**Methods:** From 2016 to 2018, 350 faecal specimens were obtained from patients with an HIV/AIDS positive status associated with diarrhoea attending inpatient (hospitalisation) and outpatient care units of El Hadi Flici (ex El- Kettar) hospital, Alger city, Algeria, and screened for the presence of *Cryptosporidium* using microscopy. Positive samples were submitted to the “Centre National de Référence-Laboratoire Expert-Cryptosporidioses”, Rouen University Hospital, France, for molecular analysis (species, genotype) by DNA sequencing of the *SSU18S rRNA* and *Gp60* genes, respectively.

**Results:** Out of 350 samples, 33 (9.4%) were microscopically positive for *Cryptosporidium* spp. of which 22 isolates were successfully amplified at the 18S rRNA and gp60 loci. Based on sequence analysis: 15 isolates were identified as *C. parvum* with family subtypes IIa-7, and IId-8, while 5 were identified as *C. hominis* (family subtypes Ia-2 and Ib-3) and 2 as *C. felis*.

**Conclusion:** The predominance of *C. parvum* subtype families IIa and IId in this study highlights the potential importance of zoonotic cryptosporidiosis transmission to Algerian HIV-positive subjects. More extensive sampling of both humans and farm animals, especially sheep, goats and calves, and collection of epidemiological data are needed for better understanding of the sources of human *C. parvum* infections in Algeria.

**Author summary:** Cryptosporidiosis, an opportunistic infection, still represents a severe threat for HIV-infected individuals. *Cryptosporidium parvum* and *Cryptosporidium hominis* are the leading cause of human cryptosporidiosis. Besides, other species and genotypes of *Cryptosporidium* might infect both immunocompetent and immunocompromised subjects.

In Algeria, no study has been conducted until now on the prevalence and molecular characteristics of *Cryptosporidium*-infection among HIV-infected individuals. Thus, this study aimed to examine the distribution and molecular characteristics of *Cryptosporidium* spp—isolates to provide clues to the understanding of transmission dynamics of species and genotypes to Algerian HIV-infected patients.

Of 350 faeces samples, 33 were microscopy-positive for *Cryptosporidium* and molecular characterisation obtained for 22 isolates resulted in the identification of *C. hominis, C. parvum*, and *C. felis*. The frequent occurrence of the zoonotic IIa and IId subtype families of C. parvum was suggestive of widespread zoonotic transmission of cryptosporidiosis in Algeria, and warrants further extensive molecular epidemiological studies in both human and animal populations.

## INTRODUCTION

Human Immunodeficiency Virus (HIV)-infection continues to be a significant global public health issue nowadays. End 2018, 37.9 million people were globally living with HIV/ Acquired Immune Deficiency Syndrome (AIDS) [1]. In the Middle East and North Africa (MENA) region, an extensive geographic area that extends from Morocco to Iran (encompassing approximately 22 countries), the estimated prevalence rate of adults aged15-49 years living with HIV infection is one of the lowest in the world (less than 0.1%). In Algeria, available data indicate 11 000 to14 000 people living with HIV, among whom 68–82% had access to antiretroviral therapy [2].

Diarrhoea is a common clinical manifestation of HIV infection regardless of whether patients have AIDS. Indeed, one of the hardest-hit organs in HIV infected individuals is the intestinal tract. Enterocytic-HIV infection results in enterocyte atrophy and impaired functioning, destruction of gut immune cells and intestinal dysfunction are resulting in diarrhoea [3,4].

Diarrhoea is a significant cause of morbidity in HIV patients, and nearly 40% of those who die of AIDS experienced diarrhoea [5,6]. In HIV patients, cryptosporidiosis is an opportunistic infection and an indicator of full symptomatic AIDS [7] and was reported as the leading indicator of death in adult Kenyan patients [6,8,9].

Despite the availability of antiretroviral therapy (with unequal access worldwide), prevalence rates of cryptosporidiosis remain high among HIV-infected patients, as illustrated by values of 26.9 and 26.7% in Ethiopia and Iran, respectively [10,11]. In Morocco, a neighbouring country, although no data are available concerning the incidence of cryptosporidiosis in HIV-infected patients, 2 respiratory cryptosporidiosis cases were reported in this population under high active antiretroviral therapy (HAART) [12]. Therapeutic intervention leads to recovery of the CD4+T cells count in HIV/AIDS patients [13]. In a murine model, resolution of established *Cryptosporidium parvum* infection requires CD4+Tcells and gamma interferon [14]. In HIV patients, CD4+T cells count <100 cells/mm^3^ were associated with susceptibility to the *Cryptosporidium* infection [15]. CD4+T cells count are useful in predicting the course of *Cryptosporidium* infection as in many other opportunistic infections [16].

Based on biological and molecular characteristics, 31 different *Cryptosporidium* species have been currently identified, while many other genotypes are still of uncertain taxonomic status [17,18]. With the development of molecular epidemiology, more and more data are available worldwide, enabling better knowledge of *Cryptosporidium* spp. distribution and especially its zoonotic versus anthroponotic transmission. Oocysts have been found in the faeces of many vertebrates, including domestic bovines, ovines, caprines and birds [18,19].

Regarding human infection, many species of *Cryptosporidium* have already been isolated in infected patients worldwide, i.e. the widely predominant *C. hominis* and *C. parvum*, and *C. canis, C. felis, C. meleagridis, C. muris, C. andersoni, C. cuniculus*, a *Cryptosporidium* rabbit genotype, a *Cryptosporidium* cervine genotype, and *C. serpentis* birds [20]. Regardless of immune status, the *C. hominis* species has been reported to be the predominant species infecting humans with an anthroponotic transmission in many studies, including in Africa [20]. *C. parvum* appears to be a human-adapted zoonotic species with a possible person to person transmission [21]. In addition to *C. parvum* and *C. hominis, C. meleagridis* infections are also relatively frequent in humans. In Africa, this species is more frequently implicated in immunocompromised populations (Up to 21% vs 10% in immunocompetent subjects) [20]. In Algeria, although the availability of information about the distribution of *Cryptosporidium* species in livestock (sheep and goats) [18, 22, 23], no epidemiologic report is currently available for human cryptosporidiosis. This study was aimed at providing the first description of the distribution of *Cryptosporidium* species and subtypes in a group of well-defined in Algerian HIV-infected patients.

## PATIENTS AND METHODS

### Patients, faeces sampling and microscopy

From 2016 to 2018, faecal specimens were obtained from 350 patients with an HIV/AIDS positive status associated with diarrhoea attending inpatient (hospitalisation) and outpatient care units at El Hadi Flici (ex El-Kettar) hospital Alger city, Algeria. After informed consent, patients filled a comprehensive questionnaire with items on age, sex, contact with animals, (pets and farm animals) and sources of drinking water. Clinical characteristics, including diarrhoea, weight loss, vomiting, abdominal pain and nausea, types of HAART drug regimens (1st line, second line and third-line therapies) are detailed in table 1. Laboratory characteristics, including blood CD4+ T-cell count, were recorded by physicians in charge. Cryptosporidium microscopy-based screening was performed in El Hadi Flici Ex El-Kettar hospital, Alger city, Algeria. Faecal specimens were examined by direct microscopy before and after concentration using a modification of the methods described by Ritchie [24]. All specimens were smeared onto glass slides, stained using the modified Ziehl Nielsen and auramine techniques [25] and examined using light (1,000 X) and fluorescence (100 X and 400 X) microscopy respectively. A sample was considered *Cryptosporidium*-positive if typical oocysts of 4–6um diameter were visible. Positive samples were transferred to the Centre National de Référence - Laboratoire expert -crypyosporidioses (CNR-LE) (Rouen University hospital, France) for molecular analysis.

**Table 1:** Socio-demographic and clinical characteristics of HIV infected patients with microscopy positive for *Cryptosporidium* spp.

| Sex | Age intervals (years) | Duration of diarrhoea | Subtypes | CD4 (cells/mm3) | ART regimen | contact with animals | Water consumption |
| --- | --- | --- | --- | --- | --- | --- | --- |
| F | 50-60 | > 14days | IlaA15G2R1 | 26 | virologic failure | Cats | Tap water |
| M | 60-70 | > 14days | IIdA19G1 | 45 | New case | Sheep and cattle | Well water |
| F | NA | > 14days | IIdA19G1 | 20 | New case | No | Tap water |
| F | 40-50 | > 14days | - | 74 | lost to follow-up | Cats and turtle | Tap water |
| M | 20-30 | > 14days | *C.felis* | 57 | New case | No | Well water |
| M | 10-20 | > 14days | *C.felis* | NA | New case | Cats and pigeons | Tap water |
| M | 40-50 | > 14days | IIdA19G1 | 1 | New case | No | Well water |
| M | 40-50 | > 14days | IIdA16G1 | 172 | 3TC/DRV/RTV | No | Tap water |
| F | 30-40 | NA | - | 106 | New case | No | Tap water |
| M | 40-50 | > 14days | - | NA | 3TC/EFV/AZT | Sheep and cattle | NA |
| F | 80-90 | <14days | IlaA21G1R1 | 512 | 3TC/EFV/ABC | NA | Tap water |
| F | 30-40 | NA | - | 100 | New case | NA | NA |
| M | 30-40 | > 14days | - | 45 | 3TC/EFV/ABC | No | NA |
| F | NA | NA | IIdA19G1 | NA | New case | No | NA |
| M | 50-60 | > 14days | IlaA20G1R1 | 55 | New case | NA | Tap water |
| F | 40-50 | > 14days | IIdA16G1 | 16 | New case | NA | NA |
| F | 50-60 | > 14days | IlaA16G2R1 | 40 | 3TC/DRV/RTV | NA | Tap water |
| M | 30-40 | > 14days | IlaA15G2R1 | 178 | 3TC/EFV/ddI | NA | Tap water |
| M | 40-50 | > 14days | IIdA16G1 | 92 | New case | Sheep and cattle (Sheep breeder) | Well water |
| M | NA | > 14days | IlaA15G2R1 | 207 | 3TC/DRV/RTV | NA | NA |
| M | 60-70 | <14days | IaA14 | NA | New case | Living in rural areas of farmed livestock | Tap water |
| F | 50-60 | > 14days | - | 112 | 3TC/EFV/AZT | Cats | NA |
| F | 30-40 | > 14days | IbA10G2 | 9 | 3TC/DRV/RTV | No | Bottled water |
| M | 20-30 | > 14days | - | 11 | New case | No | NA |
| F | 30-40 | > 14days | - | 15 | New case | No | NA |
| F | 20-30 | > 14days | IlaA14G2R1 | 50 | New case | No | Tap water |
| F | 20-30 | > 14days | IbA13G3 | 109 | New case | No | Tap water |
| F | NA | > 14days | IbA13G3 | 47 | 3TC/EFV/ABC | No | Tap water |
| M | <10 | > 14days | IaA22R2 | 7 | 3TC/EFV/AZT | No | Tap water |
| F | 20-30 | > 14days | - | 111 | New case | NA | Tap water |
| M | 40-50 | > 14days | IIdA16G1 | 53 | New case | Sheep and cattle (Sheep breeder) | Well water |
| M | 20-30 | > 14days | - | 50 | New case | NA | NA |
| M | 20-30 | > 14days | - | 48 | 3TC/EFV/ABC | 3TC/EFV/ABC | Tap water |

Abbreviations:
F: female; M: male
NA: not available
ART regimen: Lamivudine (3TC); Efavirenz (EFV); Abacavir (ABC); Didanosine (ddI); Zidovudine (AZT) ; Darunavir (DRV) ; Ritonavir (RTV).
New case: no information was available during the investigation period as sampling was performed prior to starting antiviral treatment.

### Genetic Cryptosporidium characterisation

#### DNA extraction

DNA was extracted using the QIAamp PowerFecal DNA Kit (Qiagen, France) according to the manufacturer’s recommended procedures. DNA was stored at −20°C until analysis.

#### 18s rRNA-based Cryptosporidium species identification

*Cryptosporidium* species were screened using 18S rRNA gene real-time PCR, as described elsewhere [26]. Briefly, PCR was carried out in duplicates and consisted of two duplex reactions: (i) a genus-specific PCR amplifying ∼300 bp of the *Cryptosporidium 18S rRNA* gene, duplexed with a *C. parvum*-specific PCR amplifying 166 bp of the LIB13 locus, and (ii) a *C. hominis*-specific PCR amplifying 169 bp of the LIB13 locus. Thermocycling conditions were as follows: 95°C for 10 min, followed by 55 cycles of 95°C for 15 s and 60°C for 60 s. Data were collected from each probe channel during each 60°C annealing/extension phase. Alongside real-time PCR, genomic DNAs were subjected to PCR-based sequencing of *18s rRNA* as described elsewhere [27]. A two-step nested PCR protocol was used to amplify the *18S rRNA* gene (215bp). For primary PCR, the cycling protocol was: 94°C for 5 min (initial denaturation), followed by 30 cycles of 94°C for 45 s (denaturation), 45°C for 2 min (annealing) and 72°C for 1.5 min (extension), with a final extension of 72°C for 10 min. For secondary PCR, the protocol was: 94°C for 5 min, followed by 35 cycles of 94°C for 30s, 55°C for 30s and 72°C for 30s, with a final extension of 72°C for 10 min. *C. hominis, C. parvum*, and no-template PCR controls were included in each run for each protocol.

#### Gp60 sequence amplification

Genotyping was performed by sequencing a fragment of the *Gp60* gene. Primers AL3531 and AL3533 were used in the primary PCR and primers AL3532 and LX0029 in the secondary PCR leading to amplification of a fragment of approximately 364 bp [28]. Each PCR mixture (total volume, 50 μl) contained 5 μl of 10X DreamTaq Buffer, each deoxynucleoside triphosphate at a concentration of 0.2 mM, each primer at a concentration of 100 nM, 2.5 U of DreamTaq polymerase, and 5µL of DNA template. Also, 1.25µL of DMSO (100%) was added to the mixture. A total of 40 cycles, each consisting of 94°C for 45 s, 55°C for 45 s, and 72°C for 1 min, were performed. An initial hot start at 94°C for 3 min and a final extension step at 72°C for 7 min was also included. Each amplification run included a negative control (PCR water) and two positive controls (genomic DNA from *C. parvum* oocysts purchased from Waterborne Inc., and *C. hominis* genomic DNA from a faecal specimen collected in Rouen University Hospital). Products were visualised in 2% agarose gels using ethidium bromide staining, and identification was confirmed by sequencing. Positive samples were further genotyped by DNA sequencing of the *Gp60* gene amplified by a nested PCR following the protocol described elsewhere [28].

#### DNA sequence analysis

Sequencing was used to confirm *Cryptosporidium* species/genotypes from second-round PCR products. PCR amplicons were purified using Exonuclease I/Shrimp Alkaline Phosphatase (Exo-SAP-IT) (USB Corporation, Cleveland, USA). They were sequenced in both directions using the same PCR primers at 3.2 uM in 10 μl reactions, Big Dye™ chemistries, in ABI 3500 sequence analyser (Applied Biosystems, CA, USA). Sequence chromatograms of each strand were examined with 4peaks software and compared with published sequences in the GenBank database using BLAST (www.ncbi.nlm.nih.gov/BLAST).

#### Consent and ethical approval

The authors confirm that all the participants were apprised about the aims of the study protocol. Those aged <18 years, consents were obtained from parents or guardians. Participants were also informed of the right to refuse to participate or withdraw from the study at any time without giving any reason. This study was approved by the Ethical clearance committee of the El Hadi Flici Ex El-Kettar hospital.

#### Statistical analysis

The results obtained were presented using tables and charts (descriptive statistics). Using R statistical software (version 3.6.3), Chi-square and Fisher’s exact tests were used to check for an association between *Cryptosporidium* and factors studied. Values of *p < 0.05* were considered statistically significant.

## RESULTS

### Clinical characteristics of patients

Of individual faecal samples from 350 HIV patients examined for the presence of *Cryptosporidium* oocysts, 33 (15 female and 17 male patients) were found positive. The median age of these patients was 40 years (range 7-82 years). Reported cases were highest among patients aged 20-50 years (figure1). The major clinical symptoms consisted of watery diarrhoea in all patients (chronic in 32, intermittently in one) which might be associated with nausea, vomiting or abdominal pain (n=32). Besides, fever, asthenia and weight loss were reported in 8, 16 and 23 patients respectively. Less frequent, headache or cognitive impairment was associated with Cryptosporidium infection (n=5). Mean and median values of CD4+ cell counts were 81.65 cells/mm^3^ and 50 cells/mm3 (range 1-512 cells/mm^3^) respectively. Correlation of the *Cryptosporidium* infection in HIV patients with their CD4+ cell count proved that the patient with CD4 count of <100 cells/mm^3^ were 6.36 times more likely to have the *Cryptosporidium* infections with a *p-value <0.001*.

**Figure 1.**
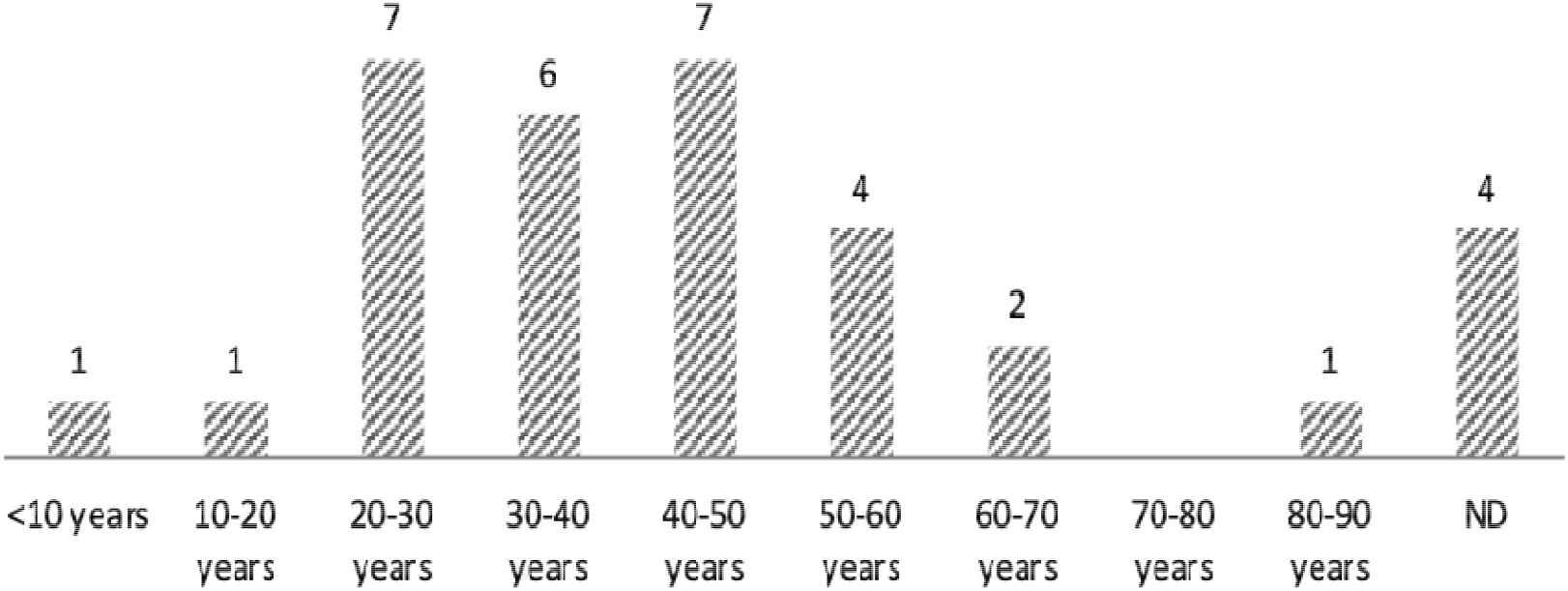
Age distribution of *Cryptosporidium*–infected HIV/AIDS patients.

### *Cryptosporidium* species and *gp60* genotypes distribution

Twenty-two of the 33 positive isolates were successfully amplified at the 18S rRNA and gp60 locus. Based on sequence analysis: *C. parvum* was identified in 15 samples with family subtypes IIa (n=7), and IId (n=8). *C. hominis* was detected in 5 cases (family subtypes Ia (n=2) and Ib(n=3) and 2 patients were infected with *C. felis*. Heterogeneity of *Cryptosporidium* was observed, eleven subtypes were identified, including 7 C. parvum subtypes (IIaA14G2R1, IIaA15G2R1, IIaA16G2R1, IIaA20G1R1, IIaA21G1R1, IIdA16G1 and IIdA19G1); and 4 *C. hominis* subtypes (IaA24, IaA22R2, IbA10G2 and IbA13G3). Among *C. parvum* subtypes, IIdA16G1 and IIdA19G1 had the highest occurrence followed by IIaA15G2R1. For *C. hominis* isolates, IbA13G3 was identified in 2 specimens and the other subtypes in one sample each (Table I). Unique sequences generated in this study were deposited in GenBank under accession numbers MT084775-MT084794.

### Association between treatment status and Cryptosporidium spp infection

Among the 22 patients with GP60 characterised *Cryptosporidium* spp infection, 9 documented-patient reported adherence to HAART (Table 1) and distributed as follows: 1°) Four patients initiated first-line ART regimen consisting of a combination between nucleoside analogue reverse transcriptase inhibitors (NRTIs) and non-NRTIs. Lamivudine (3TC) and Efavirenz (EFV) was commonly used as the backbone in first-line therapy. HAART regimen was diverse: 3TC/EFV/Abacavir (ABC) (n=2); 3TC/EFV/Didanosine (ddI) (n=1); and 3TC/EFV/Zidovudine (AZT) (n=1). The results of the *Gp60* subtyping showed one C. hominis Ib family (IbA13G3); and within C. parvum, 2 subtype family IIa (IIaA15G2R1 and IIaA21G1R1). 2°) Four individuals were using the second-line regimen. The favoured second-line therapy was a double boosted protease inhibitor combination regimen consisting of Darunavir (DRV) boosted with Ritonavir (RTV) in association with 3TC. Subtype IbA10G2; IIaA15G2R1; IIaA16G2R1 and IIdA16G1 were detected. 3°) C. parvum IIaA15G2R1 was identified in a patient with virologic failure on second-line ART regimen. Virologic failure represents the definition of viral non-suppression (plasma HIV RNA > 1000 copies/mL) used by the WHO Public health approach for low-and middle-income countries. As for whether it was the first or the second-line regimen, no significant association was found between *Cryptosporidium* infection and HAART treatment at the species and subtype levels

## DISCUSSION

Cryptosporidiosis is a significant cause of chronic diarrhoea and death in HIV/AIDS patients [30]. Diarrhoea occurs in 90% HIV/AIDS patients in developing countries and about 30–60% in developed countries [31,32]. Cryptosporidiosis, one of the conditions which according to the CDC classifications defines AIDS in adults and adolescents [33] and Category C: Severely symptomatic in children, is a significant cause of chronic diarrhoea in HIV/AIDS patients [34]. Of the 33 HIV-positive patients infected with characterised *Cryptosporidium* spp. in this study, thirty-one patients reported persistent chronic diarrhoea (>14 days).

In industrialised nations, access to HAART has significantly reduced the morbidity and mortality of cryptosporidiosis [35]. Algeria has provided HAART free of charge since 1998: standing out as one of the countries in the MENA region with the most advanced health responses. In 2016, the prevalence of HIV infected people in Algeria was about 0.1% [13 000 –15 000 individuals]. The results of the present study show a prevalence of cryptosporidiosis of 9.42% (33/350) among HIV/AIDS patients. In Tunisia, a neighbouring country, 42/526 included outpatients and inpatients presented *Cryptosporidium* spp oocysts in faeces. Of the 42 positive cases, six were found in HIV/AIDS patients [36]. Higher infection rates were reported among African HIV/AIDS patients such as in Ethiopia, Kenya, Nigeria, South Africa and Uganda with 26.9, 34, 22, 24.8 and 73.6% respectively [10,30,37-39].

Although there is a reduced risk of opportunistic infections in HIV-infected patients on HAART, opportunistic gastrointestinal infections may still occur. *Cryptosporidium* spp. the infection has been reported in patients with advanced immunodeficiency who are on HAART, which might explain their dyspeptic symptoms [40].

A CD4+ cell count below 50 is associated with severe disease. We found a mean CD4+ cell count of 81.65±98.36 cells/mm^3^ and a median of 50; which is consistent with the findings of others. Despite eradication report of *Cryptosporidium* spp. infection among immunocompromised patient [35,41] and the excellent virological and immunological response with an increased CD4 absolute number over time with the use of double boosted-PI regimen plus 3TC as second-line treatment [42], in the current study, patients were not able to clear off the infection, and their CD4 counts remained below 200 cells/mm3 which aligns with previous studies [43,44].

The emergence of drug-resistant HIV variants and failure or discontinuation of HAART (as the effectiveness of HAART highly depends on how adherent patients are on their treatment), the emergence of the re-emergence of *Cryptosporidium* spp. infection in these patients should be seriously considered [45,46].

To our knowledge, this is the first study of the distribution of *Cryptosporidium* species and subtypes in HIV/AIDS patients in Algeria. *C parvum* was the most common species responsible for cryptosporidiosis, followed by *C. hominis* and *C. felis*. In immunocompromised people, *C. hominis* is the most dominant species reported in Australia, Thailand, South Africa, Portugal and Peru [47]. A high diversity of *C. parvum* subtypes was observed in this study. Our results show that infections were marked by zoonotic isolates of *C. parvum* (subtypes IIa and IId), suggesting that animal-to-human transmission may be a standard transmission route of *Cryptosporidium* in Algeria.

In the IIa family subtype, the most prevalent subtype corresponds to IIaA15G2R1 (n=3/15). This subtype is the most dominant subtype infecting especially dairy cattle and has been widely reported in zoonotic infection [29,48]. As a risk factor for human cryptosporidiosis, contact with cattle or consumption of raw milk was suggested to be implicated in neighbouring countries as Tunisia [49]. Interestingly, in Algeria, this subtype has never been reported in cattle or other animals. More investigations should be performed with more substantial and more representative cattle samples in the country.

The IId family is generally considered as sheep and goat subtype, even if it has already been identified in human [50,51]. Subtype IIdA16G1 (n=4/15) identified in this study was recently reported in Algerian sheep [52]. Subtype IIdA19G1 (n=4) was also detected, which previously had been reported in goats in Spain [53], in both HIV-positive patients and pre-weaned dairy cattle in China [54,55] but had never been reported in goats or other animals in Algeria. Analysis of questionnaire answers showed that 3 of 8 patients harbouring subtype IId reported, (i) contact with animals or their excreta (living in rural areas of farmed livestock and working as a sheep breeder), and (ii) consumption of well water, a truck driver infected with *C. parvum* IIdA19G1 also noted drinking well water on his journey south. The CD4+ cell count of 6 out of 8 HIV/AIDS patients harbouring family subtype IId was under 100 cells/mm^3^.

Sequences analysis of *C. hominis* isolated subtypes showed the presence of IaA14, IaA22R2, IbA10G2 and IbA13G3. The IbA13G3 subtype is rarely isolated in human, but imported cases of cryptosporidiosis have already been reported in Spain [56] in HIV-positive individuals from Peru, Nigeria and Cameroon [57,58].

Potential zoonotic transmission to *C. felis* (n=2) was highlighted in this study. *C. felis* usually affects cats; a patient infected with this species reported, in the questionnaire, close contact with cats and birds. In Africa, reports of human infection with *C. felis* are scarce. Still, *C. felis* has been reported in HIV patients in Ethiopia [10], in HIV and non-HIV infected patients in Nigeria [43], and children under 5 years in Kenya [9]. Anthroponotic transmission of *C. felis* can occur in HIV patients, particularly in areas with a high incidence of cryptosporidiosis [59].

In the present study, we have documented the occurrence of *Cryptosporidium* infection in HIV/AIDS patients in Algeria and the characterisation of *Cryptosporidium* subtypes. Not only the findings generated from this study improve our understanding of molecular epidemiology of cryptosporidiosis in Algeria, but they contribute to the mapping of the epidemiology of *Cryptosporidium* in the MENA region too. The predominance of the *C. parvum* family subtypes IIa and IId in this study highlights the potential role and the importance of animals in the transmission pathway of human cryptosporidiosis. However, more extensive sampling of both humans and farm animals, especially sheep, goats and calves, and collection of epidemiological data are needed for a better understanding of the sources of *C. parvum* infections in human in Algeria.

## Data Availability

sequences generated during the study are deposited and available at https://www.ncbi.nlm.nih.gov/Genbank under GenBank accession MT084775- MT084794

## ACKNOWLEDGMENTS

The authors are grateful to Nikki Sabourin-Gibbs, Rouen University Hospital, for her help in editing the manuscript.

## AUTHORS CONTRIBUTIONS

Conceived and designed the experiments: SM, LF, AHH, RR. Performed the experiments: SM, AN,CM, DC, VV, RR. Analyzed the data: AM, JJB, LF, RR. Contributed reagents/materials/analysis tools: SM AHH Wrote the paper: SM, JJB, LF, RR.

